# Multi-Omics-Based Sex Stratification Identifies Distinct High-Risk Phenotypes in HFpEF

**DOI:** 10.64898/2026.07.09.26357645

**Authors:** Ekaterina E. Esenkova, Thomas Koeck, Steffen Rapp, Katrin I. Bauer, Silav Zeid, Felix S. Rausch, Philipp S. Wild, Elena Casiraghi, Elisa Araldi

## Abstract

**Background:** Heart failure with preserved ejection fraction (HFpEF) accounts for more than half of heart failure cases and is characterized by substantial clinical and biological heterogeneity. Sex differences are central to HFpEF pathophysiology, yet current phenotyping approaches often aggregate women and men, potentially obscuring distinct molecular mechanisms of disease progression. Molecularly resolved, sex-specific stratification is therefore needed to identify divergent risk pathways and improve biological understanding of HFpEF heterogeneity.

**Methods:** In 698 HFpEF participants from the prospective MyoVasc cohort (379 females, 319 males), we run separate analyses on sex-specific cohorts. For each cohort, we integrated 92 circulating proteins (Olink Inflammation panel) and 49 clinical variables using Similarity Network Fusion to construct sex-stratified patient-patient similarity networks. Spectral clustering identified sex-specific prognostic subgroups related to the primary endpoint, i.e. worsening of Heart Failure (WHF). XGBoost models characterizing cluster-defining features were validated in an independent cohort of 342 HFpEF patients from the Gutenberg Health Study (GHS; 194 females, 148 males).

**Results:** Two clusters emerged in each sex, with high-risk and low-risk clusters, showing the difference in WHF risk (MyoVasc females: HR 2.45, 95% CI 1.32-4.54, p=0.005; males: HR 2.77, 95% CI 1.27-6.04, p=0.011; C-index 0.62-0.63). Kaplan-Meier analyses confirmed separation (p<0.02 females, p<0.01 males). Clusters were reproduced in GHS using MyoVasc-trained XGBoost (females p=0.0082, males p=0.037). Shared top-ranking features included VEGF-A, TNFRSF9, and TGF-α. Females were characterized by inflammatory (CD40, HGF, TNF) and glycemic signatures, whereas males showed prominence of immune-regulatory markers (IL-10RB, PD-L1) and renal function indicators (eGFR, creatinine).

**Conclusions:** Sex-stratified molecular-clinical networks define prognostically distinct HFpEF subgroups with robust external validation. Shared protein biomarkers alongside sex-specific drivers reveal complementary progression mechanisms, supporting precision medicine strategies targeting high-risk cluster patients in sex-specific manner.

**Graphical Abstract of the Study:** 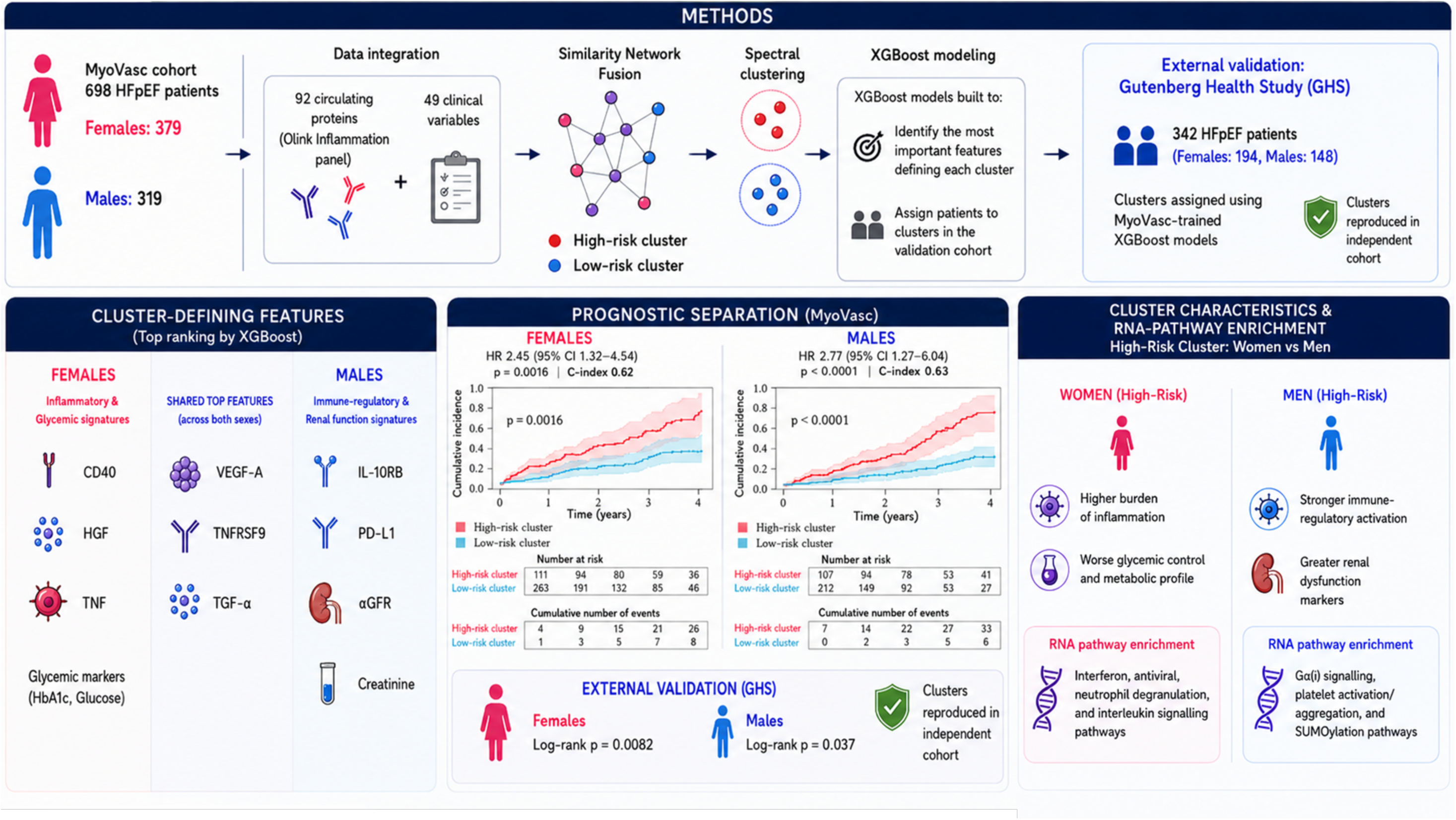

## Introduction

Heart failure with preserved ejection fraction (HFpEF) is a clinically and biologically heterogeneous syndrome and accounts for more than half of all heart failure cases (1). It is increasingly recognized as a systemic syndrome driven by a constellation of proinflammatory comorbidities, including obesity, type 2 diabetes, hypertension, chronic kidney disease, and atrial fibrillation (1). These conditions promote a chronic inflammatory milieu that contributes to coronary microvascular endothelial dysfunction, oxidative stress, and disrupted cellular protein homeostasis, ultimately leading to myocardial structural and functional abnormalities (1).

A defining feature of HFpEF is also its pronounced sex disparity (2): women are disproportionately affected, present with distinct comorbidity profiles, and often exhibit different clinical manifestations and outcomes compared with men. A 2024 Heart Failure Association of the ESC scientific statement emphasized that women with heart failure (HF) differ from men in disease presentation, diagnostic pathways, comorbidity burden, and treatment patterns, arguing that sex-specific mechanisms remain insufficiently incorporated into HF care (3). In obesity-related HFpEF, an analysis of the STEP-HFpEF trials showed that semaglutide reduced weight in women to a greater extent than men, with mechanism of it being still unclear(4).

Despite these differences, current diagnostic and therapeutic approaches remain largely sex-agnostic, contributing to limited treatment efficacy. A systematic review of 34 European Society of Cardiology (ESC) guidelines by Lashkarinia et al. (5) found inconsistent consideration of female-specific factors, with a predominant focus on pregnancy-related conditions, whereas menopause is largely overlooked. Besides that, a majority of large cohort studies has primarily relied on clinical phenotyping or retrospective analyses of electronic health records, which lack molecular resolution and are insufficient to capture the biological complexity of HFpEF (6–9). Consequently, molecular pathways that may differentially contribute to HFpEF in females and males remain poorly characterized.

Molecular profiling offers an opportunity to move beyond conventional clinical classification by capturing systemic inflammatory, metabolic, vascular, and tissue-remodelling processes that contribute to HFpEF pathophysiology. Integrating circulating proteomic data with detailed clinical phenotypes may therefore enable the identification of biologically coherent patient subgroups with distinct prognostic relevance. Importantly, performing such analyses separately in women and men may reveal molecular risk pathways that would otherwise be obscured in aggregated cohorts, opening potential therapeutic avenues.

In this study, we applied a sex-stratified, multi-omics, network-based approach in deeply phenotyped individuals with HFpEF. Proteomic and clinical phenomic data were integrated separately in women and men using Similarity Network Fusion (10) to construct sex-specific patient similarity networks (PSN) and identify molecular clusters. We then characterized these clusters according to worsening heart failure risk, evaluated cluster-discriminating features using interpretable machine learning models with validation in an independent cohort, and explored underlying biological pathways using bulk RNA-seq enrichment analysis. Through this framework, we aimed to uncover sex-specific molecular HFpEF phenotypes and identify biological pathways associated with differential disease progression in women and men.

## Methods

### Study and validation cohorts

Data were derived from the MyoVasc study (NCT04064450, https://www.clinicaltrials.gov/, registration date 2019-08-09), an investigator-initiated, single-center, prospective cohort study (11) conducted at the University Medical Center Mainz, Germany. The study was designed to investigate the natural history and progression of the HF syndrome and its risk factors. At the baseline visit, all participants underwent a standardized five-hour examination in the dedicated study center. This included comprehensive clinical and cardiovascular phenotyping, imaging procedures, cardiopulmonary tests, and laboratory measurements, together with the collection of plasma, serum, DNA/RNA, and cellular samples (11). Participants are re-examined on-site every two years to allow longitudinal assessment. The primary endpoint of MyoVasc is worsening of HF, defined as HF-related hospitalization or cardiac death. For the present analysis, only baseline data were used. Clinical HF was defined according to the 2021 Universal Definition and Classification of Heart Failure (12), as a clinical syndrome characterized by symptoms and/or signs caused by a structural and/or functional cardiac abnormality and corroborated by elevated natriuretic peptide levels and/or objective evidence of pulmonary or systemic congestion. Accordingly, the MyoVasc HF definition and phenotypic classification used in the present analysis follow the 2021 consensus framework by Bozkurt et al (12).

For external validation, we used a subsample of HFpEF participants from the Gutenberg Health Study (GHS). GHS is a large, population-based, prospective cohort study from the Rhine-Main region in Germany, designed to investigate cardiovascular risk factors and outcomes in the general population (13). In the present work, we included GHS participants fulfilling diagnostic criteria for HFpEF and with available phenotypic and protein data comparable to MyoVasc, thereby enabling validation of the identified clusters in an independent cohort. All participants of MyoVasc (reference number 837.319.12 (8420-F)) and GHS (reference number 837.020.07(5555)) provided written informed consent, and the study was approved by the local ethics committee of the University Medical Center Mainz in accordance with the Declaration of Helsinki (11). All study procedures adhered to the principles of Good Clinical and Epidemiological Practice.

### Data collection and acquisition

All biospecimens were processed and collected under standardized pre-analytical conditions and stored at −80 °C within two hours of collection at the BBM Biobank in Mainz, Germany. Blood was drawn into PAXgene Blood RNA Tubes (BD, Becton, Dickinson and Company, Franklin Lakes, NJ, USA). Once-thawed EDTA-anticoagulated blood plasma samples were analyzed with proximity extension assay technology (Olink Biosciences, Uppsala, Sweden), a targeted method of protein expression quantification that produces normalized expression values (14). Specifically, the Inflammation Olink panel was used for the present analysis, covering 92 proteins. The panel includes cytokines, chemokines, growth factors, immune-cell activation markers, receptors, and other proteins involved in inflammatory signaling, leukocyte recruitment, endothelial activation, tissue remodeling, and immune regulation.

RNA isolation was performed in an automated manner using the Maxwell® RSC system in combination with the Maxwell® RSC simplyRNA Blood Kit (both Promega Corporation, Madison, WI, USA). For transcriptome profiling, 3′ mRNA sequencing was conducted. Library preparation was carried out at the NGS Core Facility of the Institute of Human Genetics, Genomics Core Unit, Life & Brain Research Center, University Hospital Bonn (Bonn, Germany). Libraries were generated using the QuantSeq 3′ mRNA-Seq Library Prep Kit (Lexogen GmbH, Vienna, Austria), which produces Illumina-compatible libraries enriched for sequences near the 3′ end of polyadenylated transcripts (a more detailed description in Supplementary Methods).

Clinical data comprised routine blood measurements, anthropometric variables, blood pressure measurements, cardiac biomarkers, and echocardiographic parameters. Routine laboratory measurements included electrolytes, renal function markers, liver and enzyme markers, inflammatory markers, glycemic markers, lipid markers, thyroid-stimulating hormone, hematological indices, and differential blood counts. Anthropometric and hemodynamic measurements included age, body weight, height, waist and hip circumference, waist-to-hip ratio, systolic blood pressure, and diastolic blood pressure. Echocardiographic parameters included E/e′ ratio as a surrogate measure of left ventricular filling pressure, left ventricular ejection fraction, left ventricular mass, and relative wall thickness. Echocardiographic acquisition and quantification were performed using highly standardized protocols and according to the 2015 recommendations of the American Society of Echocardiography and the European Association of Cardiovascular Imaging for cardiac chamber quantification (15). A complete list of all clinical parameters is provided in Supplementary Table 3.

### Definition of worsening of heart failure

The primary endpoint of the MyoVasc study was worsening of heart failure (WHF). It was defined as a composite endpoint comprising (i) heart failure–related hospitalization, or (ii) cardiac death. Heart failure hospitalizations were defined as unplanned inpatient admissions with a primary diagnosis of heart failure requiring intravenous diuretic therapy. Cardiovascular death included death due to progressive heart failure, sudden cardiac death, or death of presumed cardiovascular origin. All events were ascertained through systematic review of hospital records, outpatient documentation, and death certificates, and were adjudicated according to predefined study criteria.

### Data Handling and Statistical Analysis

We focused on individuals with HF with preserved ejection fraction (HFpEF) from the MyoVasc cohort (11). Before combining the protein and clinical data, we processed them to handle missing values and reduce redundancy. Missing data were imputed, using missForest R package (16). We also removed variables that were either highly similar to others (pairwise correlation cutoff >=0.9) or showed almost no variation (findCorrelation and nearZeroVar functions in caret R package), to avoid biasing the results (17). For sex-specific analyses, the study population was stratified into female and male subgroups. All subsequent analyses were performed separately for each sex.

To integrate the protein and clinical data, we applied Similarity Network Fusion (SNF) (5), an integrative approach that combines multiple data modalities by constructing patient–patient similarity networks for each data type and iteratively diffusing information across them by exploiting shared neighborhood structure. A comprehensive mathematical description of the SNF methodology is provided in the Appendices. Following the construction of a fused similarity network from proteomic and phenomic data, patients were grouped into two clusters using spectral clustering. The resulting clusters were evaluated with respect to the clinical outcome of worsening of HF using age-adjusted Cox proportional hazards regression and defined as high-risk and low-risk clusters.

### Validation and molecular characterisation of clusters

After defining clusters as high- or low-risk clusters according to the Cox proportional hazard regression using worsening of HF as a primary endpoint, we characterized and validated their prognostic relevance, by applying Extreme Gradient Boosting (XGBoost), a supervised machine learning model (18). XGBoost model was trained to discriminate between high- and low-risk clusters identified in the MyoVasc discovery cohort based on molecular features, leveraging its ability to capture nonlinear relationships and complex feature interactions. Subsequently, the trained XGBoost was applied to an independent validation cohort, GHS, where model performance and generalizability with respect to WHF were evaluated. For molecular characterization, we identified the features with the highest gain values in the XGBoost model. We then compared the top-ranked features between females and males, with a particular focus on those showing sex-specific contributions.

### Pathway Enrichment Analysis of Bulk RNA-Seq Data

Variance-stabilized RNA sequencing expression data were imported and merged with clinical and cluster assignment information using unique sample identifiers. Only HFpEF participants with available RNA-seq data and corresponding cluster membership were included in the analysis. Analyses were performed in a sex-stratified manner. For each sex, individuals were assigned to previously defined high-risk and low-risk clusters derived from molecular–clinical network analysis. Gene expression values were compared between clusters using two-sided Welch’s t-tests for each gene. Log2 fold change (log₂FC) was calculated as the mean expression difference between the high-risk and low-risk clusters. P-values were adjusted for multiple testing using the Benjamini–Hochberg false discovery rate (FDR) procedure. As no individual genes remained significant after multiple-testing correction, pathway analysis was conducted using a pre-ranked GSEA approach based on the complete ranked gene list (log₂FC). This method detects coordinated pathway-level perturbations even in the absence of strongly differentially expressed single genes.

Pathway enrichment analysis was conducted using the *ReactomePA R package* (*19*) and the *gsePathway* function, implementing a pre-ranked GSEA approach based on Reactome pathway annotations. Gene sets containing fewer than 10 genes or more than 500 genes were excluded. Statistical significance was assessed using permutation-based enrichment testing, and p-values were adjusted for multiple testing using the Benjamini–Hochberg (BH) method. Pathways with an adjusted p-value < 0.05 were considered statistically significant.

## Results

### Sex-stratified network analysis identifies prognostically distinct HFpEF clusters

A total of 698 HFpEF participants from the MyoVasc cohort were included in the discovery analysis, comprising 379 females and 319 males. Using 141 variables (92 proteins, 49 clinical variables), we constructed patient-patient similarity networks and aggregated them with SNF to derive the clusters. Spectral clustering identified two clusters in each sex. To preserve statistical power and ensure stable subgroup sizes, the maximum number of clusters was prespecified as two.

Baseline characteristics of the resulting sex-specific clusters in both training and validation cohorts are provided in Supplementary Tables 1 and 2. Incidence plots revealed distinct WHF trajectories across the two clusters in the MyoVasc cohort (Fig. 1). In both females and males, one cluster, herein referred to as *high-risk cluster*, showed significantly higher cumulative incidence of worsening of HF compared to another cluster, herein referred to as *low-risk cluster* (females: p < 0.02; males: p < 0.01; Fig. 1). Age-adjusted Cox proportional hazards regression confirmed these findings for both sexes, demonstrating that individuals in the *high-risk cluster* were exposed to increased WHF risk compared to the low-risk group (females: n=379, 69 events, HR 2.45, 95% CI 1.32-4.54, p=0.005, C-index 0.63; males: n=319, 49 events, HR 2.77, 95% CI 1.27-6.04, p=0.011, C-index 0.62).

**Figure 1.**
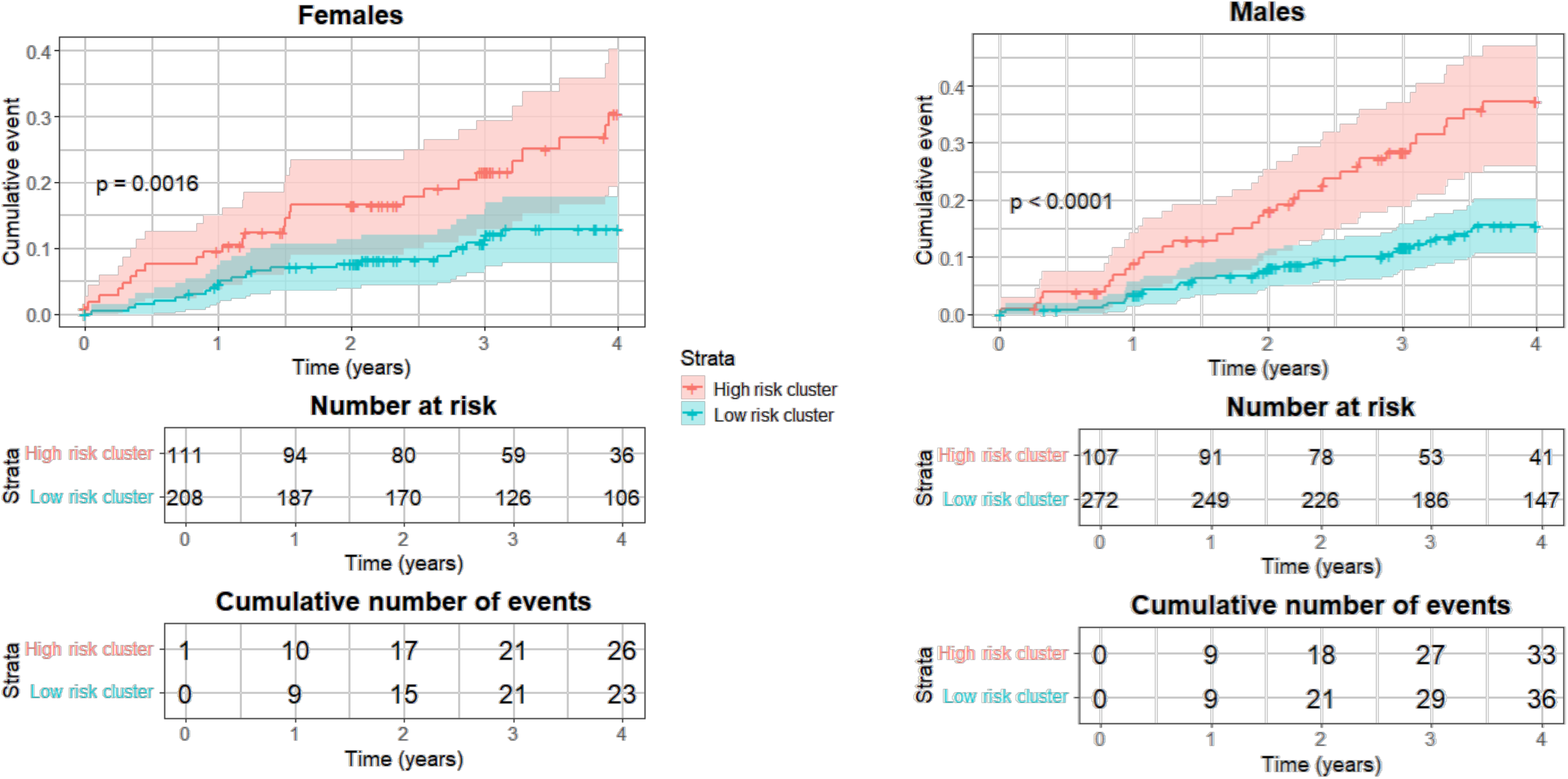
Sex-stratified cumulative incidence of worsening of heart failure (HF) across clusters in the study cohort – MyoVasc. Cumulative incidence of worsening of HF events is shown for females (left) and males (right), stratified by the two identified clusters. *High-risk cluster* (red) exhibits significantly higher WHF incidence compared to Low risk cluster (blue), with p < 0.005 in females and p < 0.0001 in males. Number at risk and cumulative events are indicated below each panel.

In both sexes, the high-risk cluster showed a greater burden of cardiometabolic comorbidities than the low-risk cluster. Among females, high-risk patients more frequently had diabetes mellitus (48.6% vs. 15.4%), obesity (55.9% vs. 35.1%), chronic kidney disease (36.0% vs. 14.4%), and cardiovascular disease (96.4% vs. 87.0%). Among males, the high-risk cluster was characterized by higher prevalence of diabetes mellitus (41.6% vs. 29.1%), atrial fibrillation (44.6% vs. 30.2%), chronic kidney disease (48.5% vs. 12.9%). The female population was predominantly postmenopausal in both risk strata, with postmenopausal women comprising 96.3% of the high-risk cluster and 88.5% of the low-risk cluster.

### Cluster prognostic value is reproduced in an independent HFpEF cohort

To evaluate the reproducibility of the identified clusters, XGBoost models were trained separately in females and males using MyoVasc cluster assignments and subsequently applied to an independent HFpEF cohort from the Gutenberg Health Study (GHS; n = 342, including 194 females and 148 males).

Patients classified into high-risk and low-risk groups in GHS showed significant differences in WHF incidence that mirrored the findings observed in MyoVasc (Figure 2). Kaplan–Meier analyses demonstrated significant separation between risk groups in both females (p = 0.0082) and males (p = 0.037), supporting the reproducibility and external validity of the identified sex-specific phenotypes.

**Figure 2.**
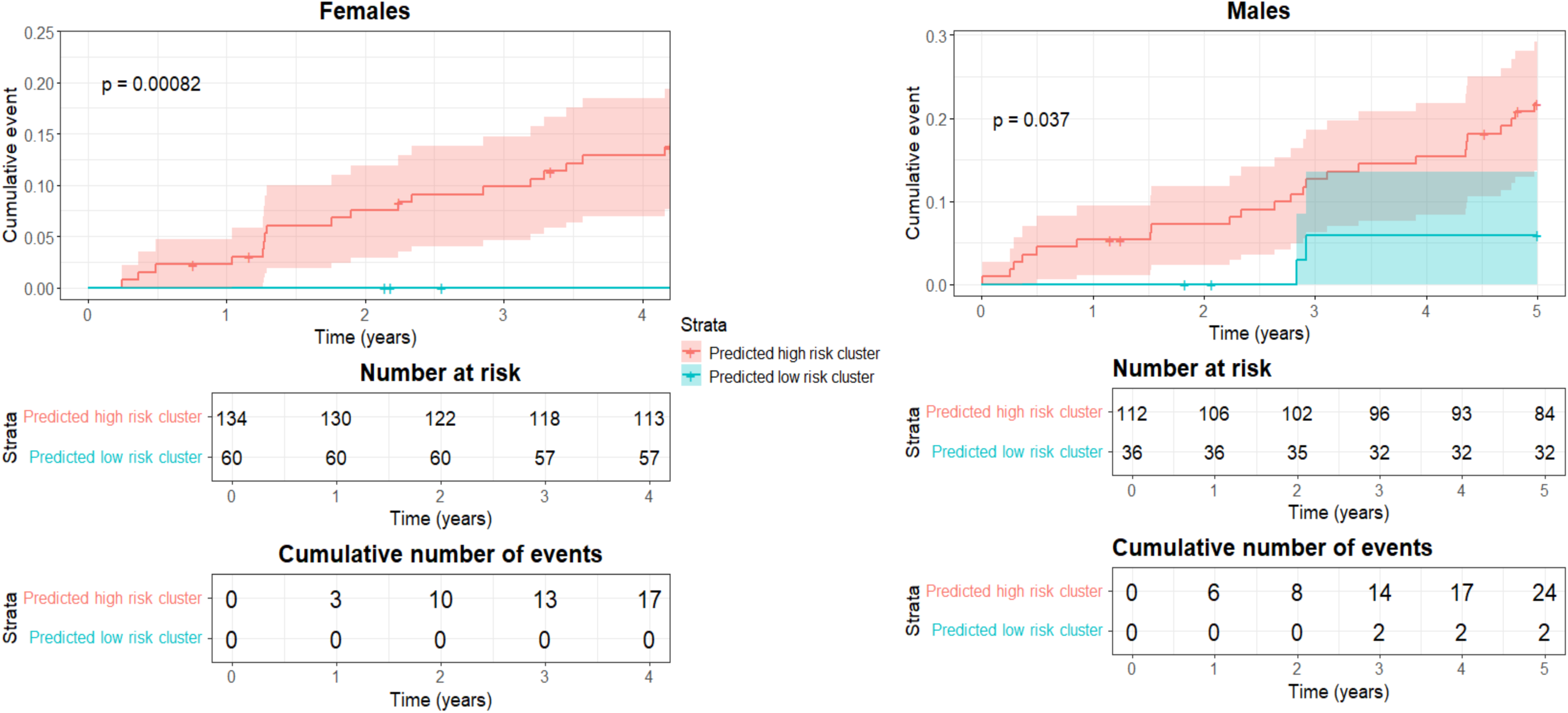
Sex-stratified cumulative incidence of worsening of heart failure (HF) by predicted risk clusters in test cohort - GHS. Cumulative incidence of worsening of heart failure (HF) is shown in females (left) and males (right), stratified by predicted risk clusters from study cohort MyoVasc (high-risk vs. low-risk) in GHS validation cohort. In both sexes, individuals assigned to the predicted high-risk cluster exhibited a higher cumulative incidence of events compared to the low-risk cluster. This difference was more pronounced in females (p = 0.00082) than in males (p = 0.037), based on log-rank tests. Shaded areas represent 95% confidence intervals.

### Shared and sex-specific molecular features characterize high-risk HFpEF phenotypes

To identify variables contributing most strongly to cluster assignment, feature importance was evaluated using sex-specific XGBoost models (Figure 3). Several proteins emerged among the highest-ranking features in both sexes, including VEGF-A, TNFRSF9, and TGF-α.

**Figure 3.**
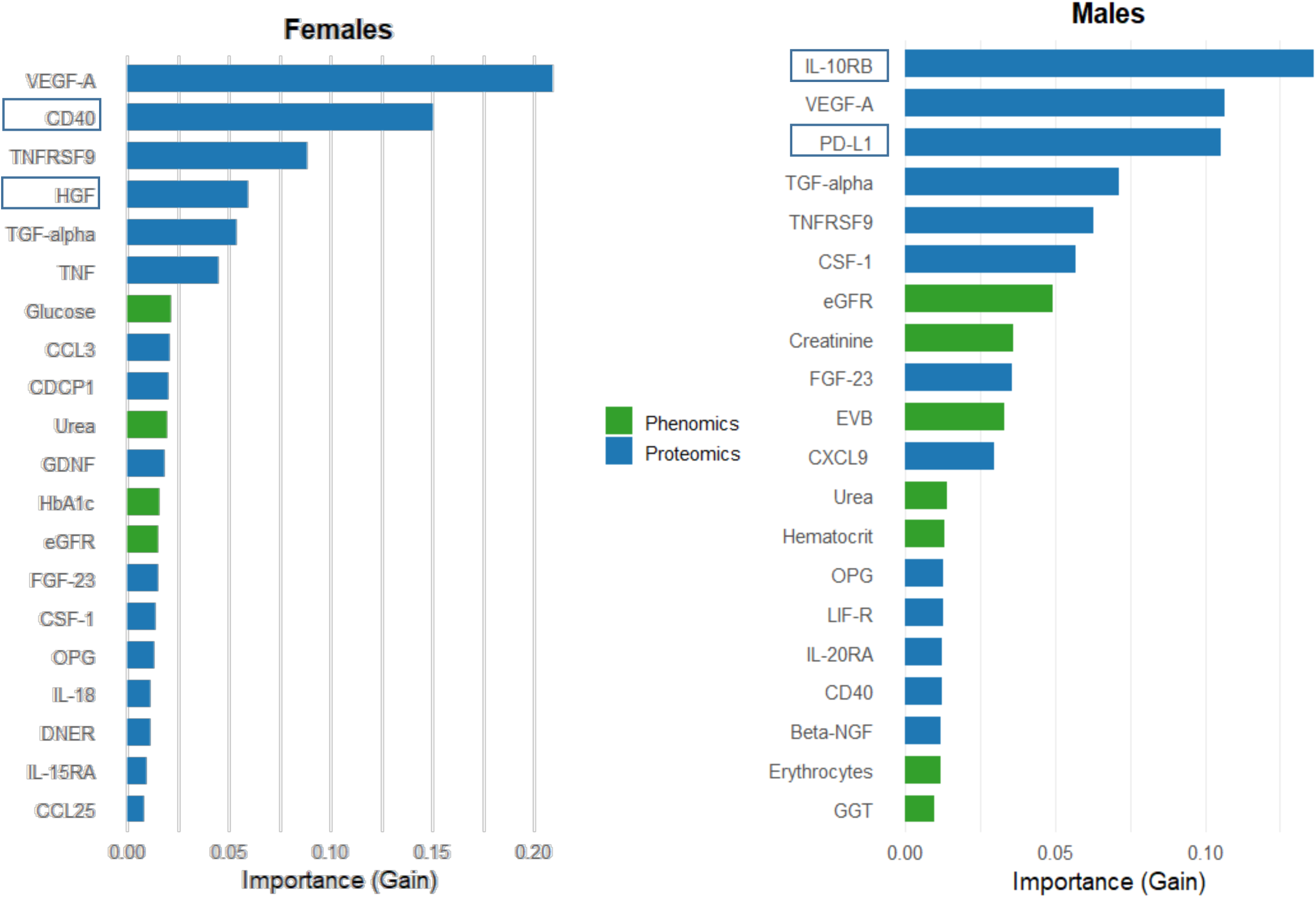
Defining features for sex-specific clusters. Barplots display the top features ranked by gain from the XGBoost model separately in females (left) and males (right). Protein biomarkers, clinical chemistry, and hematologic parameters with the highest gain values are shown, representing the variables that most strongly contribute to cluster assignment within each sex. Features marked with brackets denote sex-specific defining parameters in top features.

In females, CD40 exhibited the highest gain and was accompanied by additional inflammatory markers, including HGF and TNF, as well as glycemic parameters such as HbA1c and glucose. Consistent with the XGBoost-derived feature ranking, direct comparison of protein expression between high- and low-risk female clusters showed significantly higher levels of several top-ranked proteins in the high-risk cluster, including VEGF-A, CD40, TGF-α, TNFRSF9, HGF, FGF-23, CDCP1, TNF, GDNF, IL-15RA, CCL3, OPG, CXCL9, CSF-1, and IL10 (Fig.4). These differences were highly significant after adjustment for multiple testing, with adjusted p-values ranging from 2 × 10⁻³⁰ for VEGF-A to 6.2 × 10⁻¹⁷ for IL10. DNFR showed a smaller but significant difference between female clusters, with relatively higher expression in the low-risk cluster.

**Figure 4.**
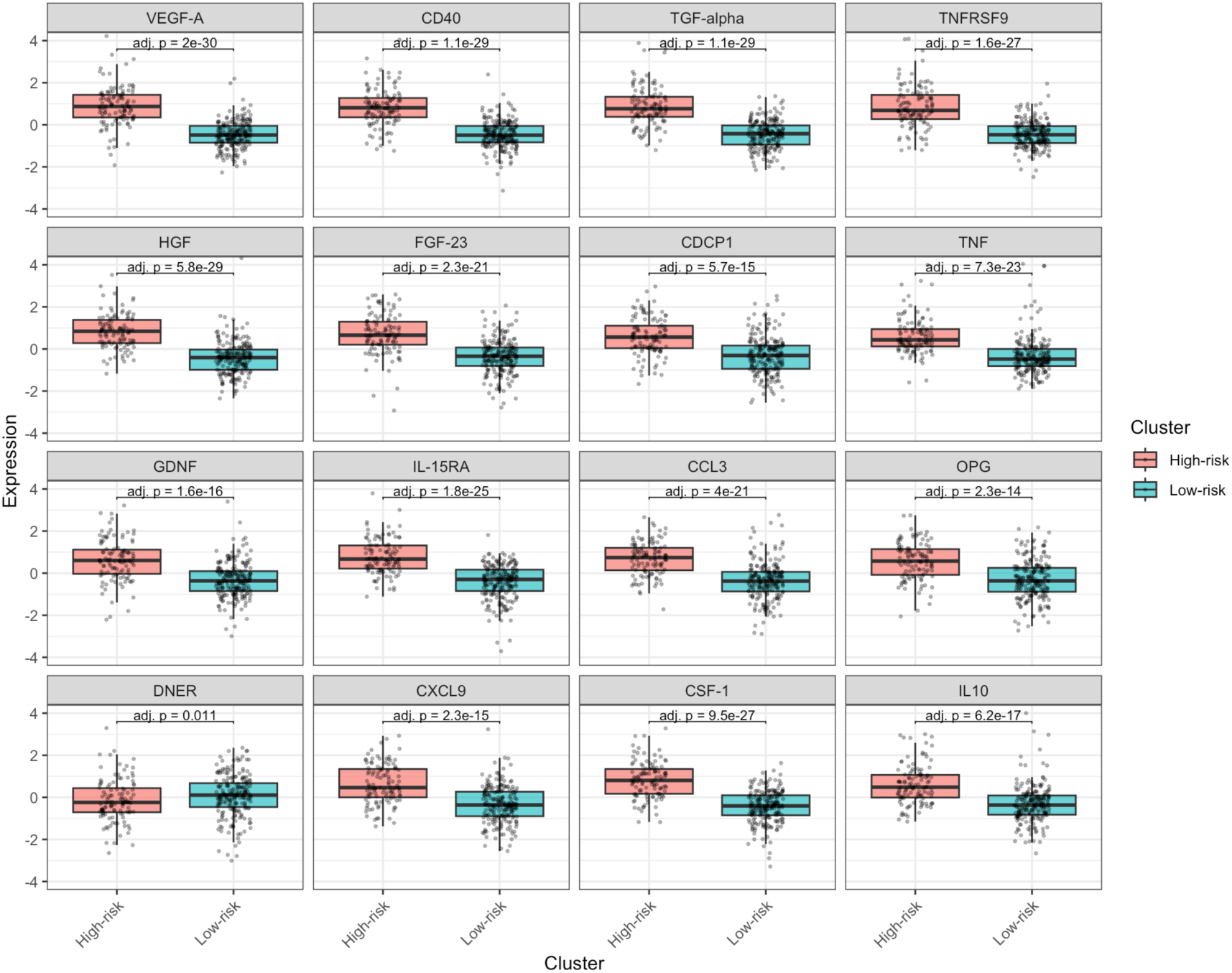
Differential protein expression between high-risk and low-risk clusters in females. Boxplots show the distribution of normalized protein expression values for the top cluster-discriminating proteins in female HFpEF patients, comparing individuals assigned to the high-risk and low-risk molecular clusters. Each panel represents one protein, with individual patient-level values shown as jittered points. Female high-risk patients showed higher expression of multiple inflammatory, immune-activation, growth-factor, and tissue-remodelling proteins, including VEGF-A, CD40, TGF-alpha, TNFRSF9, HGF, FGF-23, CDCP1, TNF, GDNF, IL-15RA, CCL3, OPG, CXCL9, CSF-1, and IL10. DNER showed a comparatively different distribution, with higher values in the low-risk group. Adjusted p-values are shown above each comparison. Boxplots indicate the median and interquartile range, with whiskers representing the distribution of values outside the interquartile range.

In males, top-ranking features additionally comprised immune-regulatory proteins such as IL-10RB, PD-L1, and CSF-1, alongside renal function markers including eGFR and creatinine. Protein-level comparisons in males likewise demonstrated a consistent increase of the top-ranked biomarkers in the high-risk cluster, including IL-10RB, PD-L1, CSF-1, TNFRSF9, VEGF-A, FGF-23, TGF-α, CXCL9, MMP-10, FGF-21, beta-NGF, SLAMF1, and IL-20RA, with adjusted p-values ranging from 2.3 × 10⁻³⁴ for VEGF-A to 7.5 × 10⁻¹⁰ for IL-20RA (Fig.5). Together, these findings indicate that the high-risk HFpEF clusters in both sexes are characterized by broad upregulation of inflammatory, immune-regulatory, growth-factor, and remodeling-associated proteins, while simultaneously revealing sex-specific molecular and clinical contributors underlying the identified high- and low-risk clusters.

**Figure 5.**
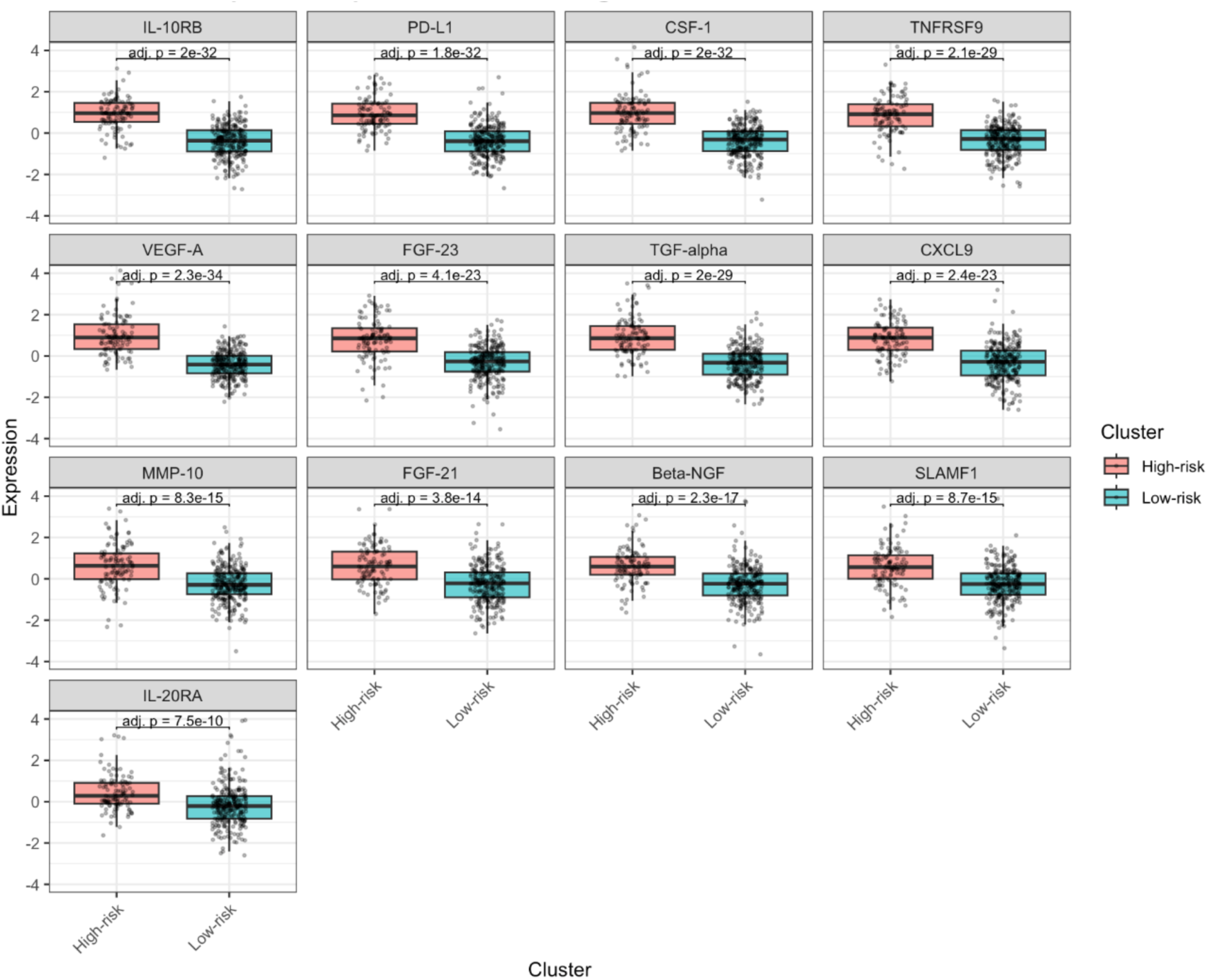
Differential protein expression between high-risk and low-risk clusters in males. Boxplots show the distribution of normalized protein expression values for the top cluster-discriminating proteins in male HFpEF patients, comparing individuals assigned to the high-risk and low-risk molecular clusters. Each panel represents one protein, with individual patient-level values shown as jittered points. Across the male cohort, high-risk patients displayed consistently higher expression of several inflammatory, immune-regulatory, growth-factor, and tissue-remodelling proteins, including IL-10RB, PD-L1, CSF-1, TNFRSF9, VEGF-A, FGF-23, TGF-alpha, CXCL9, MMP-10, FGF-21, Beta-NGF, SLAMF1, and IL-20RA. Adjusted p-values are shown above each comparison. Boxplots indicate the median and interquartile range, with whiskers representing the distribution of values outside the interquartile range.

### Transcriptomic pathway analysis reveals sex-specific biological programs

RNA sequencing data were available for a subset of HFpEF patients and were analysed separately in females and males according to cluster assignment. Although no individual genes remained significant after correction for multiple testing, preranked gene set enrichment analysis identified significant pathway-level differences between high-risk and low-risk clusters (Figure 6). In females, significantly enriched pathways were dominated by interferon- and antiviral-response programs, including interferon alpha/beta signaling, interferon gamma signaling, interferon signaling, modulation of host responses by interferon-stimulated genes, antiviral mechanisms by IFN-stimulated genes, and the ISG15 antiviral mechanism (Fig. 6). Additional enrichment of neutrophil degranulation and interleukin signaling indicated broader innate and cytokine-mediated immune activation in the female clusters.

**Figure 6.**
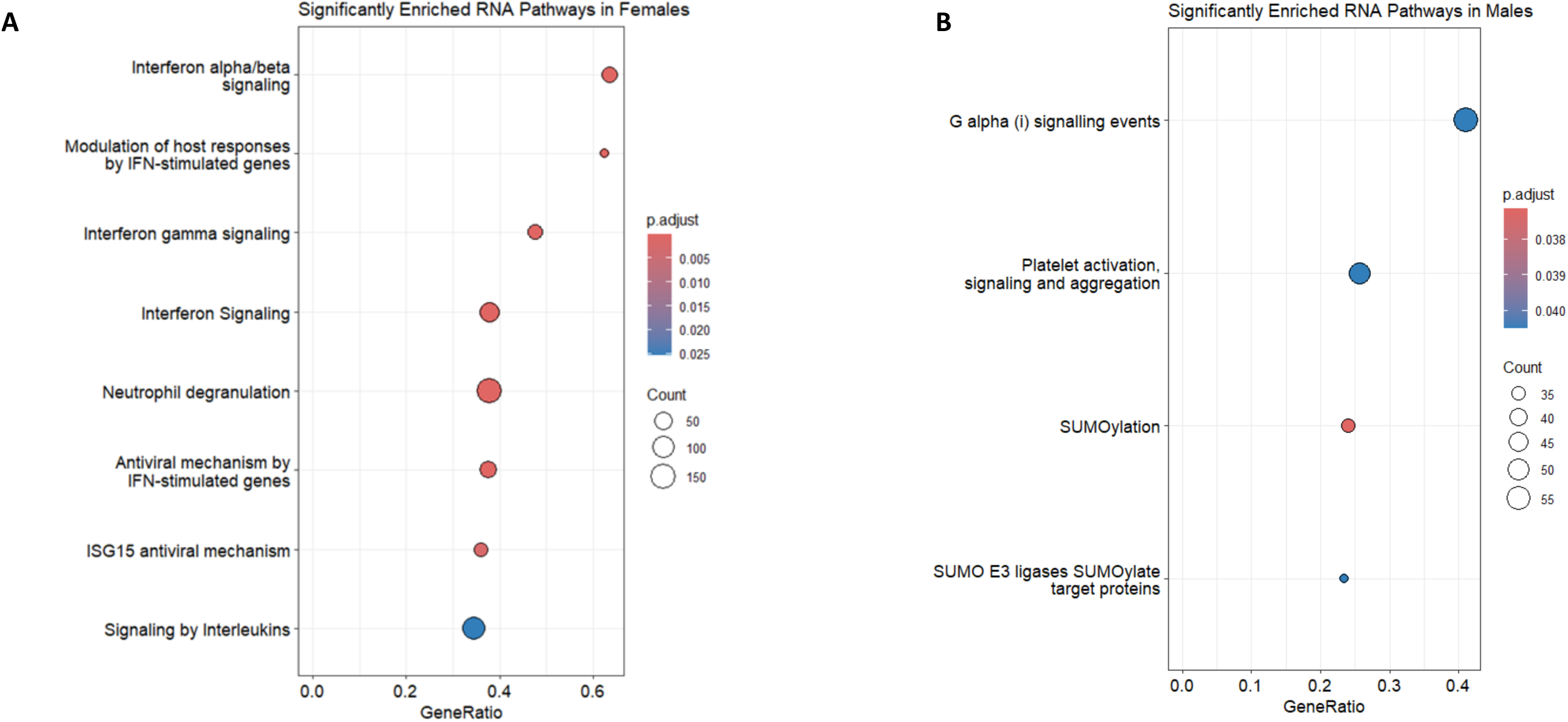
Sex-specific RNA pathway enrichment in high-risk versus low-risk clusters. Dot plots show significantly enriched RNA-derived pathways in female and male HFpEF patients. Enriched pathways are displayed separately for females (**A**) and males (**B**). The x-axis represents the gene ratio, indicating the proportion of genes from each pathway contributing to the enrichment signal. Dot size reflects the number of genes assigned to each pathway, while dot colour represents the adjusted p-value. In females, enriched RNA pathways were dominated by interferon-related, antiviral, neutrophil degranulation, and interleukin signalling programmes, indicating a pronounced innate immune and inflammatory activation signature. In males, enriched RNA pathways were mainly related to G alpha (i) signalling, platelet activation and aggregation, and SUMOylation processes, suggesting a distinct pattern involving cellular signalling, platelet biology, and post-translational regulation.

In contrast, RNA pathways enriched in males were included G alpha(i) signaling events, platelet activation, signaling and aggregation, and post-translational regulatory processes related to SUMOylation and SUMO E3 ligase activity (Fig. 6).

These findings demonstrate distinct transcriptomic pathway signatures associated with high-risk and low-risk HFpEF phenotypes in females and males.

## Discussion

### Prognostic value of the clusters

Sex-stratified patient similarity networks identified two prognostically distinct HFpEF subgroups that were reproducibly observed across the MyoVasc cohort (Fig.1) and an independent validation cohort, namely GHS (Fig.2). The consistent association of the high-risk clusters with worsening of HF across both sexes supports the robustness of the identified molecular phenotypes and highlights the value of sex-stratified multiomics integration for patient risk stratification. Importantly, despite a shared prognostic structure, the molecular determinants underlying cluster assignment differed between females and males, suggesting that similar clinical trajectories may arise through partially distinct biological mechanisms.

The coexistence of shared biomarkers together with sex-specific molecular and clinical signatures supports a model in which HFpEF progression is driven by a common disease core that is further shaped by sex-dependent inflammatory, metabolic, and organ-specific processes. These findings are consistent with the growing recognition that HFpEF represents a heterogeneous syndrome with multiple pathophysiological axes rather than a single disease entity and requires a phenotype-guided management (20–22). From a translational perspective, the identification of high-risk molecular phenotypes may help guide intensified monitoring strategies and support the future development of sex-informed therapeutic approaches.

### Shared molecular drivers of worsening heart failure across sexes

Several proteins, including VEGF-A, TGF-α, and TNFRSF9, consistently contributed to cluster assignment across both female and male models, suggesting the presence of a shared molecular framework underlying HFpEF progression (Fig.3). VEGF-A is a key proangiogenic and proinflammatory mediator that has previously been linked to myocardial infarction and adverse cardiovascular remodelling (23,24). Increased VEGF-A levels may arise in response to ischemia and contribute to vascular barrier disruption and endothelial dysfunction, mechanisms increasingly implicated in HFpEF pathophysiology. Consistent with this, accumulating evidence suggests an important role of microvascular barrier dysfunction and impaired endothelial integrity in HFpEF (25). Although studies investigating circulating VEGF-A levels in HFpEF have reported inconsistent findings (26), the recurrent identification of VEGF-A in our models supports the relevance of angiogenic and vascular remodeling pathways in worsening of HF. TGF-α, an EGFR ligand involved in inflammatory signaling and tissue remodeling, has already been associated with incident heart failure in a large multicohort proteomics study across three independent cohorts (HOMAGE, ARIC, and Framingham), supporting its potential role as a biomarker of HF progression and cardiac remodelling (27). TNFRSF9, a member of the tumor necrosis factor receptor superfamily involved in T-cell activation and immune regulation, has not previously been identified as a prominent biomarker in large heart failure cohort study (28), despite its established role in immune activation and cardiovascular inflammatory remodeling (29,30). Its emergence as a key determinant in our models may therefore indicate a potentially underexplored immune-regulatory mechanism in HF pathophysiology. Supporting this notion, epigenome-wide association analyses in patients with hypertension identified significant CpG methylation sites associated with TNFRSF9 expression, suggesting its involvement in immune-mediated vascular and cardiometabolic pathways relevant to heart failure development and progression (31).

### Female molecular drivers

Beyond this shared core, the sex-stratified analyses revealed marked biological divergence. In females, the dominant contribution of inflammatory proteins together with glycemic markers suggests a tight interplay between immune activation and metabolic dysregulation. In particular, the strong contribution of CD40-associated signaling points toward enhanced immune-endothelial interactions and vascular inflammation as central components of high-risk female phenotypes. CD40/CD40L signaling has previously been implicated in cardiovascular inflammation, endothelial dysfunction, and adverse cardiac remodeling, with elevated soluble CD40 ligand levels reported in patients with acute and chronic heart failure and associated with disease severity and cardiometabolic comorbidities (32). Experimental studies further support a role for CD40–TRAF6 signaling in myocardial macrophage and T-cell infiltration, fibrosis, hypertrophy, and adverse cardiac remodeling, reinforcing its relevance to inflammatory HF pathophysiology (33). However, despite the established role of CD40 signaling in cardiovascular inflammation, evidence regarding sex-specific CD40-associated mechanisms in HFpEF remains limited. Prior work in early menopausal women has linked menopausal symptom burden to vascular inflammation (34), including higher soluble CD40 ligand levels in women with moderate-to-severe hot flushes compared with controls, suggesting that CD40L-related inflammatory activation may be accentuated in subsets of postmenopausal women. This is particularly relevant in the present study, as the female HFpEF population was predominantly postmenopausal, with postmenopausal women comprising more than 88% of each female cluster in both the control and test cohorts. Together, these observations support the hypothesis that CD40-associated signaling may contribute to an immune–vascular inflammatory phenotype in postmenopausal women with HFpEF, although direct mechanistic evidence in HFpEF remains to be established.

In addition, the relevance of Hepatocyte Growth Factor (HGF) in our sex-specific model of HFpEF further supports activation of tissue remodelling and inflammatory repair pathways, as elevated circulating HGF levels have been associated with myocardial fibrosis, endothelial injury, and adverse outcomes in large HF and HFpEF biomarker studies (35). Notably, the same large multicohort proteomics study that identified TGF-α as a biomarker associated with incident heart failure across all three cohorts (HOMAGE, ARIC, and Framingham) also highlighted HGF among the top eight HF-associated circulating proteins (27), supporting its role as a robust biomarker linked to HF progression and cardiovascular remodelling. Previous population-based analyses additionally demonstrated that higher HGF concentrations were independently associated with incident heart failure and showed stronger associations with HFpEF than HFrEF, supporting its potential role in inflammatory and endothelial remodelling pathways particularly relevant to HFpEF development (36). Nevertheless, sex-specific associations of HGF with HFpEF phenotypes have not yet been systematically characterized. The concomitant relevance of HbA1c and glucose in our study further supports the hypothesis that cardiometabolic stress may amplify inflammatory signalling in women, contributing to disease progression through integrated immune-metabolic mechanisms. The transcriptional pathway enrichment analyses reinforce this interpretation. Female high-risk profiles were characterized predominantly by interferon-related and innate immune pathways, consistent with heightened inflammatory activation and antiviral-like immune responses (Fig.6). Collectively, these findings suggest that worsening of HF in women may be driven by coordinated activation of inflammatory, endothelial, remodelling, and metabolic pathways. This observation is consistent with prior HFpEF studies showing sex-specific circulating protein profiles and stronger links between female HFpEF biology, immune activation, inflammation, and microvascular dysfunction (37,38).

### Male molecular drivers

In contrast, males demonstrated a distinct molecular profile characterized by immune-regulatory signaling, platelet activation, and pathways involved in cellular signaling and post-translational modification (Fig. 4). Rather than broad pro-inflammatory activation, the male signatures were more consistent with regulatory and adaptive immune processes, potentially reflecting differences in myeloid cell activation or compensatory anti-inflammatory responses. Notably, IL-10RB and PD-L1 emerged among the top male-associated proteins (Fig.3), further supporting an immunoregulatory phenotype. IL-10RB is involved in anti-inflammatory cytokine signaling, whereas PD-L1 functions as a key immune checkpoint regulator that modulates T-cell activation and inflammatory responses. Their enrichment in males may therefore indicate a stronger compensatory immune-suppressive response in male HFpEF progression. The enrichment of platelet and G protein signaling pathways additionally points toward a greater contribution of vascular and thrombotic mechanisms in male disease progression (Fig.6). The prominent contribution of renal function markers further supports the concept of a stronger cardiorenal axis in males. Together with the observed signaling and platelet-related pathways, these findings suggest that worsening of HF in men may be more strongly linked to systemic vascular dysfunction, coagulation-related processes, renal impairment, and adaptive immune regulation rather than the predominantly immune-metabolic phenotype observed in females.

These findings are consistent with previous studies demonstrating sex-specific molecular profiles in HFpEF, with men showing greater associations with vascular dysfunction, extracellular remodeling, and adaptive immune signaling, whereas women more commonly exhibit inflammatory and metabolic phenotypes (37). Although IL-10RB has not been extensively studied in HFpEF specifically, immune-regulatory IL-10 signaling has been implicated in cardiac remodeling and heart failure progression, suggesting that enhanced IL-10RB-related signaling may reflect compensatory anti-inflammatory responses in male HFpEF (39). The identification of PD-L1 among the top male-associated proteins may point toward enhanced immune checkpoint regulation and adaptive immune signaling in male HFpEF (40), although the precise role of PD-L1 in HFpEF remains insufficiently characterized.

Taken together, these findings support a model in which HFpEF progression is organized around a shared inflammatory and remodeling-associated disease core, while sex-specific layers of immune regulation, metabolic dysfunction, and organ crosstalk shape individual risk trajectories. Interestingly, several of the top-ranked proteins identified in the female models have previously been implicated in HFpEF and related cardiometabolic disease processes, whereas many of the male-associated proteins remain comparatively underrepresented in the current HFpEF literature. Given the higher prevalence of HFpEF in women and the predominance of female-enriched phenotypes in prior studies, it is possible that male-specific molecular mechanisms have been comparatively under characterized. These observations underscore the importance of sex-stratified approaches in multi-omics analyses, as aggregated analyses may obscure biologically meaningful heterogeneity with potential clinical relevance. Ultimately, a more refined understanding of sex-dependent molecular phenotypes may facilitate the development of precision biomarker strategies and targeted therapeutic interventions in HFpEF.

### Strength and limitations of the current study

In this study we applied a sex-stratified patient-patient network approach using Similarity Network Fusion, allowing integration of complementary proteomic and clinical data while preserving modality-specific information. This strategy is particularly suitable for moderate-sized clinical cohorts, where conventional subgroup discovery may be limited by sample size and noise within individual data layers. By constructing and fusing patient similarity networks separately in females and males, we were able to identify biologically interpretable HFpEF subgroups with consistent prognostic relevance. The identified clusters were evaluated in two independent cohorts, MyoVasc and GHS, supporting the reproducibility of the prognostic signal. The observation that high-risk clusters were associated with worsening of HF in both cohorts and both sexes strengthen the clinical relevance of the network-derived phenotypes. In addition, the combination of SNF with interpretable machine-learning models enabled prioritization of molecular and clinical features contributing to cluster assignment, thereby linking patient stratification to plausible biological mechanisms. The study benefits from deep phenotyping, including a broad panel of circulating proteins together with clinical, metabolic, and renal markers. This allowed us to move beyond purely clinical risk stratification and investigate sex-specific molecular patterns associated with HFpEF progression.

Some limitations should also be considered. First, both cohorts predominantly comprise individuals of German origin, which may limit the generalizability of the findings to more ethnically and geographically diverse populations. External validation in larger and more diverse HFpEF cohorts will be necessary to determine whether the identified sex-specific phenotypes are broadly applicable. Second, although the clusters showed reproducible prognostic associations, the study is observational and cannot establish causal relationships between the identified biomarkers and worsening of HF. Functional studies will be required to determine whether proteins such as VEGF-A, TGF-α, TNFRSF9, CD40-associated proteins, IL-10RB, or PD-L1 actively contribute to disease progression or primarily reflect downstream disease processes. Third, residual confounding remains possible despite adjustment for relevant clinical variables. Medication use, comorbidities, treatment changes, and unmeasured lifestyle or environmental factors may have influenced both molecular profiles and clinical outcomes. Fourth, the proteomic platform captures a selected set of circulating proteins and does not fully represent the myocardial, vascular, renal, or immune-cell-specific processes that may underlie HFpEF progression. Integration with additional omics layers, including metabolomics, lipidomics, methylation, and cell-type-resolved immune profiling, may provide a more complete mechanistic interpretation. Fifth, the female HFpEF population was predominantly postmenopausal, with postmenopausal women comprising more than 88% of each female cluster in both cohorts. Therefore, the findings should primarily be interpreted in the context of postmenopausal HFpEF. At the same time, this composition reflects the epidemiology of HFpEF, which predominantly affects older and postmenopausal women, supporting the clinical relevance of the identified female high-risk phenotype. Future studies including younger women, premenopausal women, and longitudinal information on menopausal transition and hormone-related factors will be needed to clarify the extent to which these molecular signatures are driven by sex, menopause, aging, or their interaction.

## Conclusions

Sex-stratified integration of proteomic and clinical data identified reproducible HFpEF patient clusters with distinct prognostic profiles in both the MyoVasc and GHS cohorts. High-risk clusters were consistently associated with worsening of HF, indicating that network-based molecular phenotyping can capture clinically meaningful heterogeneity beyond conventional clinical characterization. Our findings suggest that HFpEF progression is structured around a shared inflammatory, angiogenic, and remodeling-associated molecular core, including proteins such as VEGF-A, TGF-α, and TNFRSF9. However, the molecular features associated with high-risk phenotypes differed between sexes. In females, worsening of HF was linked to immune-metabolic activation, CD40-associated inflammatory signaling, glycemic dysregulation, and innate immune pathway enrichment. In males, high-risk profiles were characterized by immune-regulatory markers, platelet-related signaling, renal function markers, and cardiorenal features. These results emphasize that HFpEF heterogeneity is partly sex-dependent and may be obscured in aggregated analyses. By combining Similarity Network Fusion with interpretable machine learning, this study provides a scalable framework for sex-informed precision phenotyping in moderate-sized clinical cohorts. Future validation in larger and more diverse populations, together with mechanistic studies, will be essential to determine whether these molecular phenotypes can guide biomarker development, risk stratification, and eventually personalized therapeutic strategies in HFpEF.

## Data Availability

All data in this study is under controlled access, as per the signed study participants consent. Request for access to data can be addressed to the principal investigators of the study (contact: info@myovasc).

## Availability of data and materials

This project constitutes a major scientific effort with high methodological standards and detailed guidelines for analysis and publication. The data in this study is under controlled access, as per the signed study participants’ consent. Request for access to data can be addressed to the principal investigators of the study (contact: info@myovasc).

## Code availability

Analyses were conducted using R 4.2.1. The code, used to generate the results in this paper is available on GitHub (https://github.com/ekaterinaesenkova/Multi-Omics-Based-Sex-Stratification-Identifies-Distinct-High-Risk-Phenotypes-in-HFpEF/tree/main).

## Acknowledgments

The authors express their gratitude to the study participants and the current as well as former members of the MyoVasc study team. Part of this work is included in the <u>doctoral</u> thesis of Ekaterina Esenkova.

## Authors and Affiliations

^1^Preventive Cardiology and Preventive Medicine, Department of Cardiology, University Medical Center of the Johannes Gutenberg University Mainz, Germany;

^2^German Center for Cardiovascular Research (DZHK), partner site Rhine-Main, Mainz, Germany;

^3^Clinical Epidemiology and Systems Medicine, Center for Thrombosis and Hemostasis (CTH), University Medical Center of the Johannes Gutenberg University Mainz, Germany;

^4^Department of Computer Science, University of Milan, Milan, Italy;

^5^Systems Medicine, Institute of Molecular Biology (IMB), Mainz, Germany;

^6^Environmental Genomics and Systems Biology Division, Lawrence Berkeley National Laboratory, Berkeley, CA, USA

^7^Systems Medicine Laboratory, Department of Medicine and Surgery, University of Parma, Parma, Italy.

## Authors’ contributions

EEE conceived and designed the study and the analyses, performed the analyses and interpreted the data, prepared the figures, and wrote the manuscript. TK optimized protocols, prepared the samples for OLINK proteomics, contributed to Olink data generation, pre- and post-processing. RS performed bulk RNA data preparation and generation. KIB contributed with data interpretation and literature review. SZ performed RNA data pre-processing. PSW acquired funding, conceived and designed the study, supervised the analyses and interpreted the data. EC supervised the analysis and interpreted the data. EA acquired funding, conceived and designed the study and the analyses, supervised the statistical analyses, interpreted the data, and wrote the manuscript. All authors revised the manuscript critically for important intellectual content and approved the final manuscript.

## Corresponding author

Elisa Araldi

Phone: +39 0521 902074

## Ethical approval consent to participate

The MyoVasc and GHS studies were approved by the local data protection officer and the medical association of Rhineland–Palatinate State approved (reference no. 837.319.12 (8420-F), reference number 837.020.07(5555)). All study participants provided written informed consent prior to study enrolment, and the study was conducted according to the Declaration of Helsinki, the recommendations of good clinical practice and good epidemiological practice.

## Rights and permissions

### Open Access

This article is licensed under a Creative Commons Attribution 4.0 International License, which permits use, sharing, adaptation, distribution and reproduction in any medium or format, as long as you give appropriate credit to the original author(s) and the source, provide a link to the Creative Commons licence, and indicate if changes were made. The images or other third-party material in this article are included in the article’s Creative Commons licence, unless indicated otherwise in a credit line to the material. If material is not included in the article’s Creative Commons licence and your intended use is not permitted by statutory regulation or exceeds the permitted use, you will need to obtain permission directly from the copyright holder. To view a copy of this licence, visit http://creativecommons.org/licenses/by/4

## Supplementary

**Table 1.** Baseline characteristics of female HFpEF clusters.

| Characteristics | MyoVasc (Discovery cohort) |  | GHS (validation cohort) |  |
| --- | --- | --- | --- | --- |
|  | High risk | Low risk | High risk | Low risk |
| **n** | 108 | 209 | 134 | 60 |
| **Demographics** |  |  |  |  |
| **Age [years], median (IQR)** | 73.0 (68.2 - 78.0) | 70.0 (63.4-75.0) | 68.0 (62.0-71.0) | 66.5 (61.0-70.0) |
| **Menopause status** |  |  |  |  |
| Premenopause | 3.7% (4) | 7.2% (15) | 1.5%(2) | 3.3%(2) |
| Perimenopause | 0 | 1.9% (4) | 1.5%(2) | 1.7%(1) |
| Postmenopause | 96.3%(104) | 88.5%(185) | 96.3%(129) | 88.3%(53) |
| **Cardiovascular risk factors, % (n)** |  |  |  |  |
| **Arterial hypertension** | 91.0% (101) | 86.1% (179) | 79.9% (107) | 76.7% (46) |
| **Diabetes mellitus** | 48.6% (54) | 15.4% (32) | 36.8% (49) | 3.3% (2) |
| **Dyslipidaemia** | 69.4% (77) | 65.4% (136) | 58.2% (78) | 36.7% (22) |
| **Obesity (BMI > 30)** | 55.9% (62) | 35.1% (73) | 56.7% (76) | 43.3% (26) |
| **Current smoking (incl. occasional)** | 13.5% (15) | 8.7% (18) | 11.4% (15) | 5.0% (3) |
| **Comorbidities, % (n)** |  |  |  |  |
| **Atrial fibrillation** | 36.0% (40) | 28.8% (60) | 16.8% (21) | 12.3% (7) |
| **Congestive heart failure** | 91.9% (102) | 84.1% (175) | 49.3% (66) | 36.7% (22) |
| **Coronary artery disease** | 34.2% (38) | 25.0% (52)" | 21.0% (26) | 7.4% (4) |
| **History of MI** | 17.1% (19) | 15.4% (32) | 10.8% (14) | 1.7% (1) |
| **History of stroke** | **12.6% (14)** | **8.2% (17)** | **11.2% (15)** | **1.7% (1)** |
| **History of peripheral artery disease** | 16.2% (18) | 5.8% (12) | 10.6% (14) | 10.2% (6) |
| **History of Chronic Kidney Disease** | 36.0% (40) | 14.4% (30) | 2.2% (3) | 1.7% (1) |
| **History of cancer** | 22.5% (25) | 18.8% (39) | 11.9% (16) | 15.0% (9) |

**Table 2.** Baseline charateristics of male clusters.

| Characteristics | MyoVasc (Discovery cohort) |  | GHS (validation cohort) |  |
| --- | --- | --- | --- | --- |
|  | High risk | Low risk | High risk | Low risk |
| n | 101 | 278 | 112 | 36 |
| **Demographics** |  |  |  |  |
| Age [years], median (IQR) | 73.0 (66.0 - 79.0) | 70.0 (64.0- 76.0) | 69.0(65.0- 72.0) | 65.5(57.4- 70.0) |
| **Cardiovascular risk factors, % (n)** |  |  |  |  |
| Arterial hypertension | 83.2% (84) | 87.1% (242) | 82.1% (92) | 77.8% (28) |
| Diabetes mellitus | 41.6% (42) | 29.1% (81) | 43.8% (49) | 2.8% (1) |
| Dyslipidaemia | 74.3% (75) | 87.8% (244) | 72.3% (81) | 52.8% (19) |
| Obesity | 36.6% (37) | 40.3% (112) | 57.1% (64) | 41.7% (15) |
| Smoking | 13.9% (14) | 10.8% (30) | 12.6% (14) | 31.4% (11) |
| **Medical history, % (n)** |  |  |  |  |
| Atrial fibrillation | 44.6% (45) | 30.2% (84) | 21.7% (23) | 22.9% (8) |
| Congestive heart failure | 99.0% (100) | 88.1% (245) | 51.4% (57) | 52.8% (19) |
| Coronary artery disease | 54.5% (55) | 62.9% (175) | 45.9% (45) | 13.9% (5) |
| History of MI | 32.7% (33) | 37.1% (103) | 31.5% (34) | 0% (0) |
| History of stroke | 18.8% (19) | 11.2% (31) | 10.7% (12) | 2.8% (1) |
| History of peripheral artery disease | 9.9% (10) | 9.0% (25) | 16.4% (18) | 13.9% (5) |
| History of Chronic Kidney Disease | 48.5% (49) | 12.9% (36) | 8.0% (9) | 2.8% (1) |
| History of cancer | 25.7% (26) | 18.7% (52) | 14.3% (16) | 8.6% (3) |

**Table 3.**
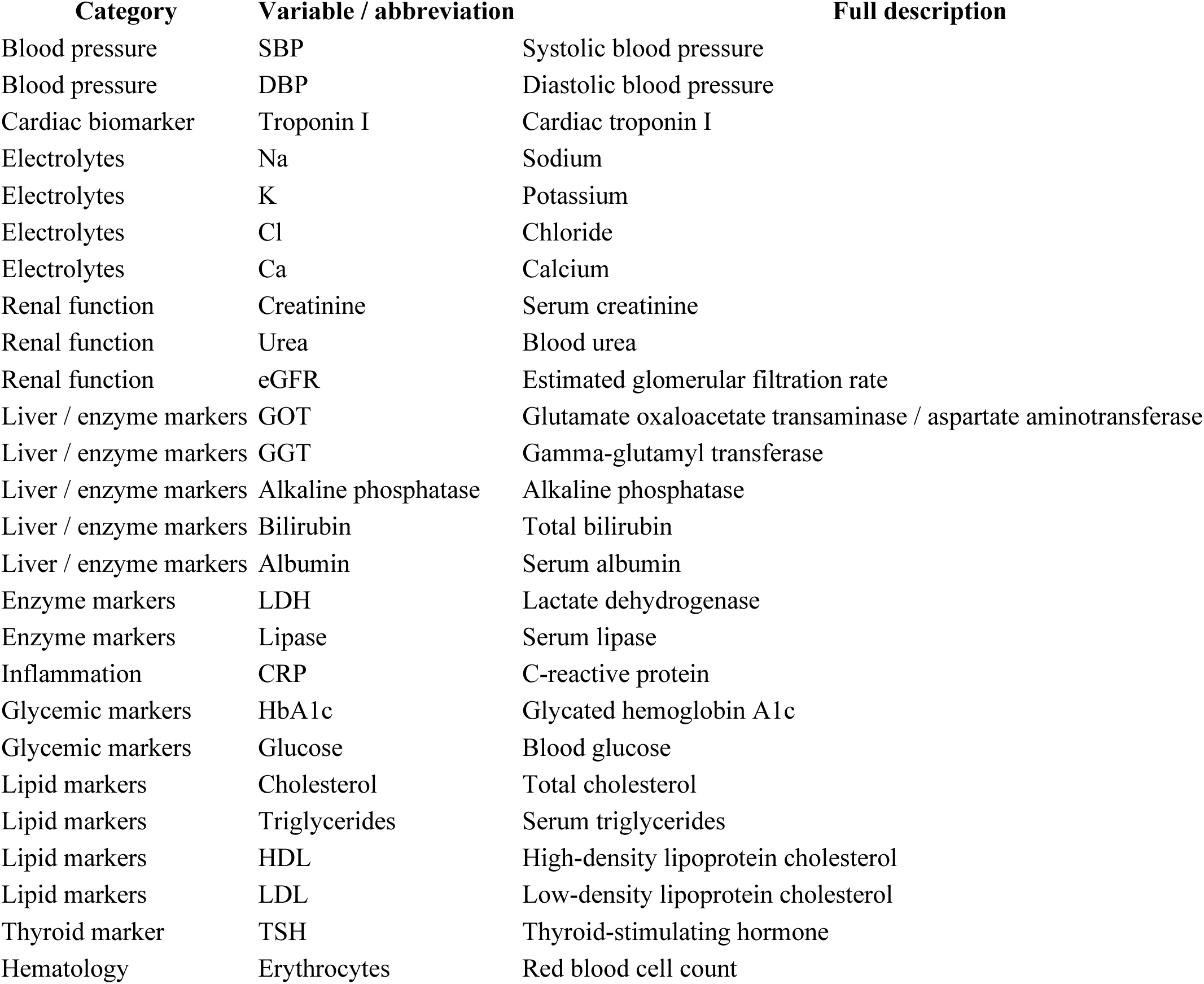
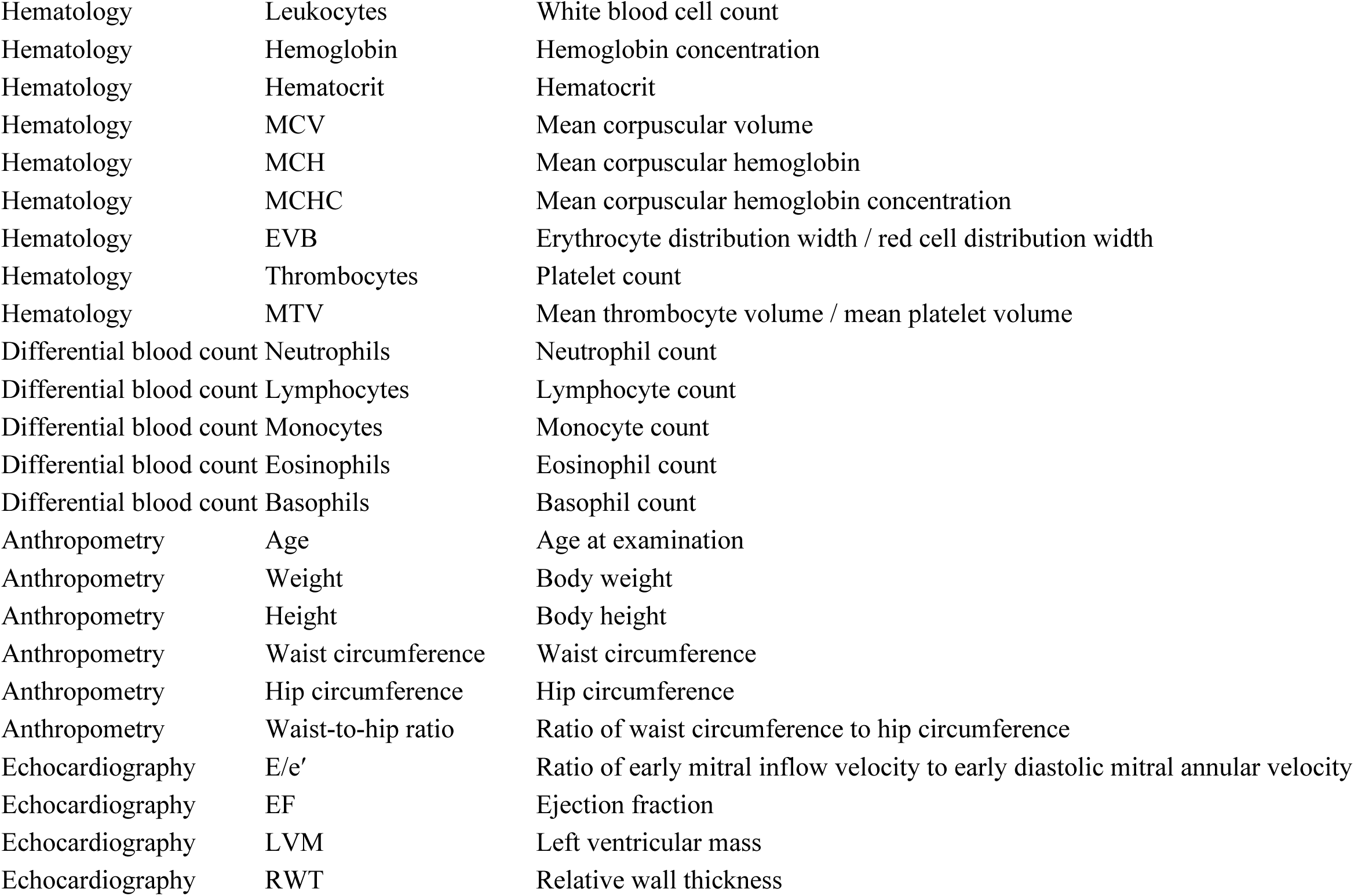
Clinical and blood routine variables included in the integrative clustering and XGBoost analyses, together with Olink Inflammation panel.

## Supplementary Methods

### Measurement and preprocessing of RNA data

Up to 200 ng of total RNA was used per sample. To mitigate the high abundance of globin transcripts in whole blood, a globin reduction add-on (Lexogen GmbH) was applied. Sequencing was performed as 51 bp single-end reads on an Illumina, Inc. HiSeq 2500 platform (San Diego, CA, USA). The target sequencing depth was approximately 10 million reads per sample, although this threshold was exceeded in some cases. Processing of raw gene-level count data was performed using the DESeq2 framework (41). To improve data quality, genes with zero expression across all samples were first removed, followed by exclusion of lowly expressed genes, defined as those with counts ≤3 in more than 60% of samples. Size factor estimation based on the median-of-ratios approach was then applied to normalize for variability in sequencing depth and RNA composition, assuming that most genes are not differentially expressed. Finally, variance-stabilizing transformation was applied to the normalized counts to reduce heteroscedasticity and enable robust downstream statistical analyses.

### Similarity Network Fusion

SNF integrates multiple data sources through a cross-diffusion process that iteratively exchanges information among the Patient Similarity Networks (PSNs) constructed from each data source (12).

More specifically, given *m* data sources, SNF first applies a scaled exponential affinity kernel to construct an individual (unimodal) PSN, represented as *W*^(*s*)^, *s* ∈ {1,…, *m*}, for each data source *s*. These PSNs are subsequently processed to generate:

1. *P*^(*s*)^, a normalized PSN capturing “global” relationships among patients; and
2. *S*^(*s*)^, a “local” PSN representing topologies within neighborhoods in *s*.

For two points *x_i_* and *x_k_*, *S*^(*s*)^(*i*, *k*) > 0 holds only if *x_k_* ∈ *N_i_*, where *N_i_* is the k-nearest neighborhood of *x_i_* in source *s*.

In the simplest case of two sources *s* ≠ *v*, the t-th iteration of the diffusion process updates 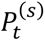 and 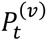 using the following equations:

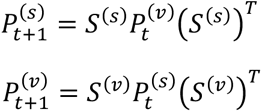

where *X^T^* is the transpose of matrix *X* Focusing on source *s* and two points *x_i_* and *x*_+_, the following equation can be expanded for a single element 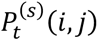 as:

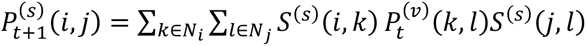

Here, the product inside the summations is non-zero only when *x_k_* ∈ *N_i_* and *x_l_* ∈ *N_j_* are also global neighbors in source *v* during the previous iteration 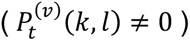. In essence, similarity information is exchanged between sources *v* and *s* only through their shared global and local neighborhoods. This iterative process progressively aligns global similarities across sources. At convergence — or after *T* iterations—the integrated (consensus) PSN, *P*^(*c*)^, is calculated as the average of *P*^(*s*)^ and *P*^(*v*)^; in other words, i.e., in the case of *m* = 2 sources, *s* and *v*:

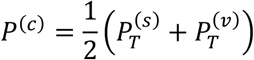

*P*^(*c*)^ captures shared and complementary global and local information from all individual data sources.

